# Firearm Purchasing and Firearm Violence in the First Months of the Coronavirus Pandemic in the United States

**DOI:** 10.1101/2020.07.02.20145508

**Authors:** Julia P. Schleimer, Christopher D. McCort, Veronica A. Pear, Aaron Shev, Elizabeth Tomsich, Rameesha Asif-Sattar, Shani Buggs, Hannah S. Laqueur, Garen J. Wintemute

**Author notes:** Corresponding author: Julia P. Schleimer, MPH.

## Abstract

**Importance:** Firearm violence is a significant public health and safety problem in the United States. A surge in firearm purchases following the onset of the coronavirus pandemic may increase rates of firearm violence.

**Objective:** To estimate the association between changes in firearm purchasing and interpersonal firearm violence during the coronavirus pandemic.

**Design:** Cross-sectional time series study. We estimate the difference between observed rates of firearm purchases and those predicted by seasonal autoregressive integrated moving average models. Using negative binomial models, we then estimate the association between excess firearm purchases and rates of interpersonal firearm violence within states, controlling for confounders.

**Setting:** The 48 contiguous states and the District of Columbia. Hawaii and Alaska are excluded due to missing or incomplete data.

**Exposure:** The difference between observed and expected rates of firearm purchases in March through May 2020, approximated by National Instant Criminal Background Check System records.

**Main Outcome and Measure:** Fatal and nonfatal injuries from interpersonal firearm violence, recorded in the Gun Violence Archive.

**Results:** We estimate that there were 2.1 million excess firearm purchases from March through May 2020—a 64.3% increase over expected volume, and an increase of 644.4 excess purchases per 100,000 population. We estimate a relative rate of death and injury from firearm violence of 1.015 (95% Confidence Interval (CI): 1.005 to 1.025) for every 100 excess purchases per 100,000, in models that incorporate variation in purchasing across states and control for effects of the pandemic common to all states. This reflects an increase of 776 fatal and nonfatal injuries (95% CI: 216 to 1,335) over the number expected had no increase in purchasing occurred.

**Conclusions and Relevance:** We find a significant increase in firearm violence in the United States associated with the coronavirus pandemic-related surge in firearm purchasing. Our findings are consistent with existing research. Firearm violence prevention strategies may be particularly important during the pandemic.

**KEY POINTS:** *Question:* Is the coronavirus-related surge in firearm purchasing associated with changes in rates of interpersonal firearm violence?

*Findings:* This cross-sectional time series study suggests the recent increase in firearm purchases—an estimated 2.1 million excess purchases nationally between March and May 2020—is associated with a statistically significant increase in firearm violence. We estimate an increase of 776 fatal and nonfatal injuries (95% CI: 216 to 1,335) in the US over the number expected for those months had there been no increase in purchasing.

*Meaning:* During the coronavirus pandemic, an acute increase in firearm access is associated with an increase in firearm violence.

---

Firearm violence is among America’s leading causes of death and disability ^1^ and has profound adverse social, psychological, and economic effects on life in this country. ^2,3^ A large body of research has established an association between the prevalence of firearm ownership and rates of both interpersonal and self-directed firearm violence at the population, ^4–6^ household, ^7,8^ and individual ^9,10^ levels. Surges in firearm purchasing, which acutely increase the prevalence of firearm ownership, have been well documented in association with mass shootings and significant political events and are followed by population-level increases in firearm violence. ^11–13^

The coronavirus pandemic has created deep and widespread social and economic disruption in the United States. As of June 30, 2020, more than 2.6 million cases and nearly 123,000 deaths have been reported. ^14^ Federal Bureau of Investigation (FBI) records of background checks pursuant to firearm purchases ^15^ suggest a substantial surge in firearm purchasing in many states beginning near the onset of the coronavirus pandemic. Given prior findings, it is reasonable to expect a subsequent increase in firearm violence.

Other effects of the pandemic, or the country’s response to it, might well modify the relationship between surges in firearm purchasing and firearm violence. Stay-at-home orders might reduce community violence, since fewer people are in public places—or increase it if fewer potential witnesses are on scene and/or law enforcement presence is reduced. The pandemic has exacerbated factors that contribute to interpersonal violence, including financial stress, tension, trauma, worry, and a sense of hopelessness. Fear and scapegoating associated with COVID-19 may increase hate crime. Violence at home might increase if stay-at-home orders increase contact between persons in violent relationships, including intimate partners, children, and vulnerable elders.

In this paper, we explore the association between trends in firearm purchasing and interpersonal firearm violence during the coronavirus pandemic, relying on publicly available data: the FBI’s National Instant Criminal Background Check System (NICS) records ^15^ as a proxy for firearm purchasing and public reports of firearm violence collected by the Gun Violence Archive (GVA). ^16^ We date the onset of the pandemic as January 21, 2020, when the virus was first reported in the US. Our period of observation extends through May 31, 2020.

## METHODS

### Design, setting, and subjects

This is a cross-sectional time series study of monthly firearm purchasing and firearm violence in the US from January 2018 through May 2020. The 48 contiguous US states and District of Columbia (DC) are included, resulting in 1,421 place-time units (29 months x 48 states and DC). Hawaii and Alaska are excluded due to missing or incomplete data.

### Data sources

We approximate firearm purchasing using monthly state-level NICS background check data ^15^ specific to firearm purchase transactions (excluding those for pawn redemptions or carry permits). Denominators for rates are obtained from the US Census’ Annual Estimates of the Resident Population for states. Although NICS checks do not have a 1:1 correspondence with purchased firearms, because most states permit multiple firearm purchases in a single transaction, this discordance is likely stable over time, leaving the data compatible with time series analyses.

GVA records of interpersonal firearm violence are based on reviews of 7,500 news outlets and other public sources. ^16^ Data used for this study include the date and location of the event and a limited set of event characteristics (Supplementary Table 1). We include only events coded as intentional, interpersonal violence with 1 or more shots fired and 1 or more persons killed or injured. We use the term ‘injuries’ to include both nonfatal injuries and deaths. GVA data have been used for research on legal intervention shootings, ^17^ firearm homicides, ^18^ mass shootings, ^19,20^ and community violence ^21^ and have performed well relative to other sources. ^17,19^

**Table 1.** State-level association between changes in firearm purchasing and interpersonal firearm violence, 2018-2020

|  | Unadjusted<sup>a</sup> |  |  | Adjusted |  |  |
| --- | --- | --- | --- | --- | --- | --- |
|  | RR | 95% CI |  | RR | 95% CI |  |
| March, 2020 | 1.001 | 0.903 | 1.109 | 0.939 | 0.861 | 1.025 |
| Cumulative excess purchases<sup>b</sup> | 1.009 | 1.000 | 1.023 | 1.015<sup>g</sup> | 1.005 | 1.025 |
| COVID-19 cases and deaths<sup>c</sup> |  |  |  | 1.018<sup>h</sup> | 1.009 | 1.028 |
| Stay-at-home order |  |  |  | 1.053 | 0.923 | 1.200 |
| Movement<sup>d</sup> |  |  |  | 1.002 | 0.999 | 1.005 |
| Average precipitation<sup>e</sup> |  |  |  | 0.998 | 0.994 | 1.003 |
| Average temperature<sup>f</sup> |  |  |  | 1.011<sup>h</sup> | 1.006 | 1.016 |

Note. Results are from negative binomial regression models with indicators for state, year, and month, and the log of the population as an offset. The outcome is counts of firearm injuries (nonfatal and fatal).
RR = rate ratio. CI = confidence interval.
<sup>a</sup> Estimates are minimally adjusted.
<sup>b</sup> Association reflects an increase of 100 purchases per 100,000 population.
<sup>c</sup> Association reflects an increase of 100 cases and deaths per 100,000 population.
<sup>d</sup> Association reflects a 1-unit increase in Apple's mobility index.
<sup>e</sup> Association reflects a 1-inch increase in average monthly precipitation.
<sup>f</sup> Association reflects a 1-degree increase in average monthly temperature, measured in degrees Fahrenheit.
<sup>g</sup> P<0.05
<sup>h</sup> P<0.001

We developed a directed acyclic graph to identify a minimum set of time-varying covariates needed to control for confounding (Supplementary Figure 1). Covariates include monthly COVID-19 cases and deaths per population, state stay-at-home orders, average monthly movement (a measure of adherence to social distancing recommendations), and average monthly temperature and precipitation. We use data on COVID-19 cases and deaths from Johns Hopkins University Center for Systems Science and Engineering. ^14^ Information on stay-at-home orders are obtained from the New York Times, ^22,23^ which maintains updated data on orders at the state level. Stay-at-home orders, which were implemented or lifted at different times, are coded as the proportion of each month that the order was in effect. Movement data are obtained from Apple’s Mobility Trends, ^24^ which compiles data on changes since January 13, 2020 in “the number of requests made to Apple Maps for directions” for various transportation types (walking, driving, and public transportation). Data on temperature and precipitation are obtained from the PRISM Climate Group at Oregon State University (Supplementary Table 2). ^25^

**Figure 1.**
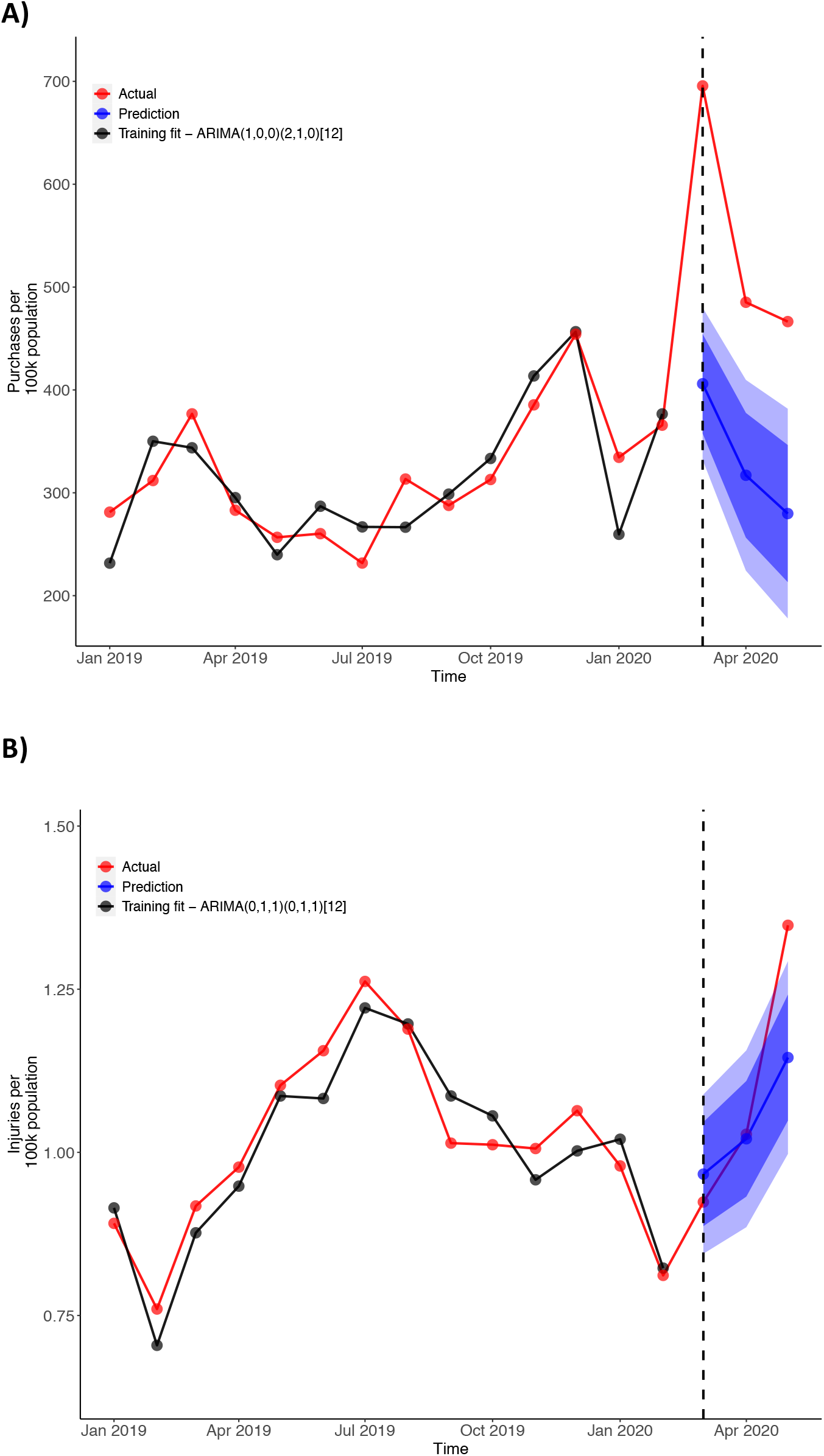
Nationwide trends in firearm purchasing (Panel A) and firearm violence (Panel B) A) Monthly firearm purchases per 100,000 population, with training data from January 2011 through February 2020. B) Monthly injuries from firearm violence per 100,000 population, with training data from January 2015 (earlier GVA data appear to reflect an undercount of events) through February 2020. Dotted line indicates March 2020. Blue bands indicate 80% and 95% prediction intervals.

### Statistical analyses

Our primary exposure is the difference between observed and expected rates of firearm purchases following the onset of the pandemic. We estimate expected rates of firearm purchases for each state in March, April, and May 2020 with seasonal auto-regressive integrated moving average (SARIMA) models, in which firearm purchasing rates are estimated as a function of prior purchasing rates (auto-regressive) and forecast errors (moving average). We fit SARIMA models to training data beginning in January 2011, so as to include prior documented spikes in firearm purchasing, ^11–13^ and ending in February 2020. Models were fit using the Hyndman and Khandakar algorithm, ^26^ and residual autocorrelation was examined using the Box-Ljung test ^27^ with the Benjamini and Hochberg ^28^ correction for multiple testing.

Changes in firearm purchasing are defined as the monthly difference between observed and expected purchasing rates, beginning in March 2020 and cumulated through May (defined as 0 in all months prior to March 2020). For example, a state with 30 excess purchases per 100,000 population in March, 20 excess purchases per population in April, and 10 excess purchases per population in May would receive a value of 30 for March, 50 for April, and 60 for May. We defined the exposure this way for two reasons. First, we are able to account for variation between months, rather than averaging change across the 3-month interval. Second, we allow for the accumulation of excess purchases over time because the risks of increased purchasing may be neither immediate nor time-limited.

We estimate the association between changes in firearm purchasing and firearm violence using multivariable unconditional negative binomial regression models, which account for overdispersion and yield unbiased estimates with fixed effects. ^29,30^ The outcome is modeled as counts of injuries (nonfatal and fatal) from interpersonal firearm violence, with the log of the population as an offset.

Models include indicators for months after the purchasing spike (1 if March 2020 or later); states to control for time-invariant characteristics of states; and year and month to control for state-invariant secular and seasonal trends, including the onset of the pandemic in January 2020. We are therefore comparing within-state changes in firearm violence between states with different magnitudes of change in pandemic-related purchasing. Models include all time-varying covariates listed above and clustered standard errors to account for within-state correlation over time. We also test a quadratic term for excess purchases to assess linearity.

In addition to modeling the accumulation of excess purchases, we conduct secondary analyses to capture delayed effects by including a one-month lag of the exposure—wherein the firearm purchasing variable is coded as 0 in all months prior to April 2020.

In exploratory analyses, we examine whether the associations differ by states’ baseline firearm ownership prevalence, socioeconomic status, racial residential segregation, urbanicity, violent crime rate, and social distancing, testing multiplicative interactions for each in separate models with alpha of 0.20. ^31^ Firearm ownership prevalence is measured by the proportion of suicides completed with a firearm. ^32,33^ Socioeconomic disadvantage is defined as the first principal component of each state’s average high school graduation rate, percentage of adults with some college education, unemployment rate, percentage of children in poverty, income inequality, and percentage of children living in single-parent households. ^34^ We use the dissimilarity index as an indicator for racial residential segregation. ^34,35^ Urbanicity is measured by the percentage of the population living in a rural area, ^36^ and violent crime by the number of offenses for murder and nonnegligent manslaughter, rape, robbery, and aggravated assault per population. ^37^ To account for compliance with distancing recommendations, we use Apple’s Mobility Index, as described above. ^24^ Additional details about each variable and data source are in Supplementary Table 2.

Because social distancing may affect where violence takes place and how many people are at risk of injury, we also model the outcome as: 1) counts of events of firearm violence (to examine changes in events independent of the number of people injured); and 2) the ratio of injuries to events. We use a negative binomial model for the former and linear model for the latter.

We test the robustness of our findings in several ways. First, we define the exposure as the cumulative percentage change in purchasing rather than the absolute change, as the magnitude of absolute change is somewhat dependent on states’ baseline purchasing rate. Second, we exclude Washington DC—which is a city, not a state—and events in which a child shot another person, as children’s intent to commit violence may be unclear. ^38^ Third, we include state-specific linear trends to adjust for unmeasured confounders that are neither time nor state-invariant. ^39^ Fourth, to test whether changes in firearm violence predate changes in firearm purchasing, we include leading values of the exposure. ^39^ Finally, we add a control for changes in all-cause mortality (excluding deaths from interpersonal firearm violence and coronavirus) to capture misclassification of coronavirus deaths and broader consequences of the pandemic on population health. Data on other causes of death, such as suicide or interpersonal violence not involving firearms, are not available.

Analyses were done with the forecast package (version 8.12) in R (version 4.0.0) and in Stata (SE 15.1). All significance tests were 2-sided and used alpha 0.05 unless otherwise noted. This study was approved by the University of California, Davis Institutional Review Board.

## RESULTS

We estimate that there were 947,788 excess purchases (95% prediction interval (PI): 705,841 to 1,189,735) in March 2020, another 550,537 excess purchases (95% PI: 247,130 to 853,944) in April, and 610,852 excess purchases (95% PI: 277,314 to 944,390) in May (Figure 1a). This represents a total of 2,109,177 excess purchases (644.4 per 100,000) over the 3-month period—a 64.3% increase over expected volume. Interpersonal firearm violence also increased nationally during this period (Figure 1b), although not above expected levels in March and April. There was a substantial increase in May, with 633 excess injuries (95% PI: 180 to 1,147) that month—a 17.7% increase over expected levels.

We find substantial variability between states in the cumulative difference between observed and expected purchasing rates during the 3-month period, ranging from −2.7 (Washington DC) to 1,454.3 (New Hampshire) per 100,000 population (average 780.9) (Supplementary Figure 2). Multivariable regression models show that changes in firearm purchasing within states are significantly associated with changes in firearm violence during this period. We estimate that an increase of 100 excess purchases per 100,000 population is associated with an increase in the rate of injuries from firearm violence (Rate Ratio (RR) 1.015; 95% Confidence Interval (CI): 1.005 to 1.025) (Table 1), controlling for effects of the pandemic common to all states. Using model predictions, we estimate an increase of 776 injuries (95% CI: 216 to 1,335) across the US from March through May 2020 over what would have been expected had no increase in purchasing occurred. This represents an increase of 7.8% (95% CI: 1.7% to 13.9%) over the 3-month period. The 1-month lagged associations are similar (Supplementary Table 3). Tests of quadratic terms provide evidence for a linear relationship.

**Figure 2.**
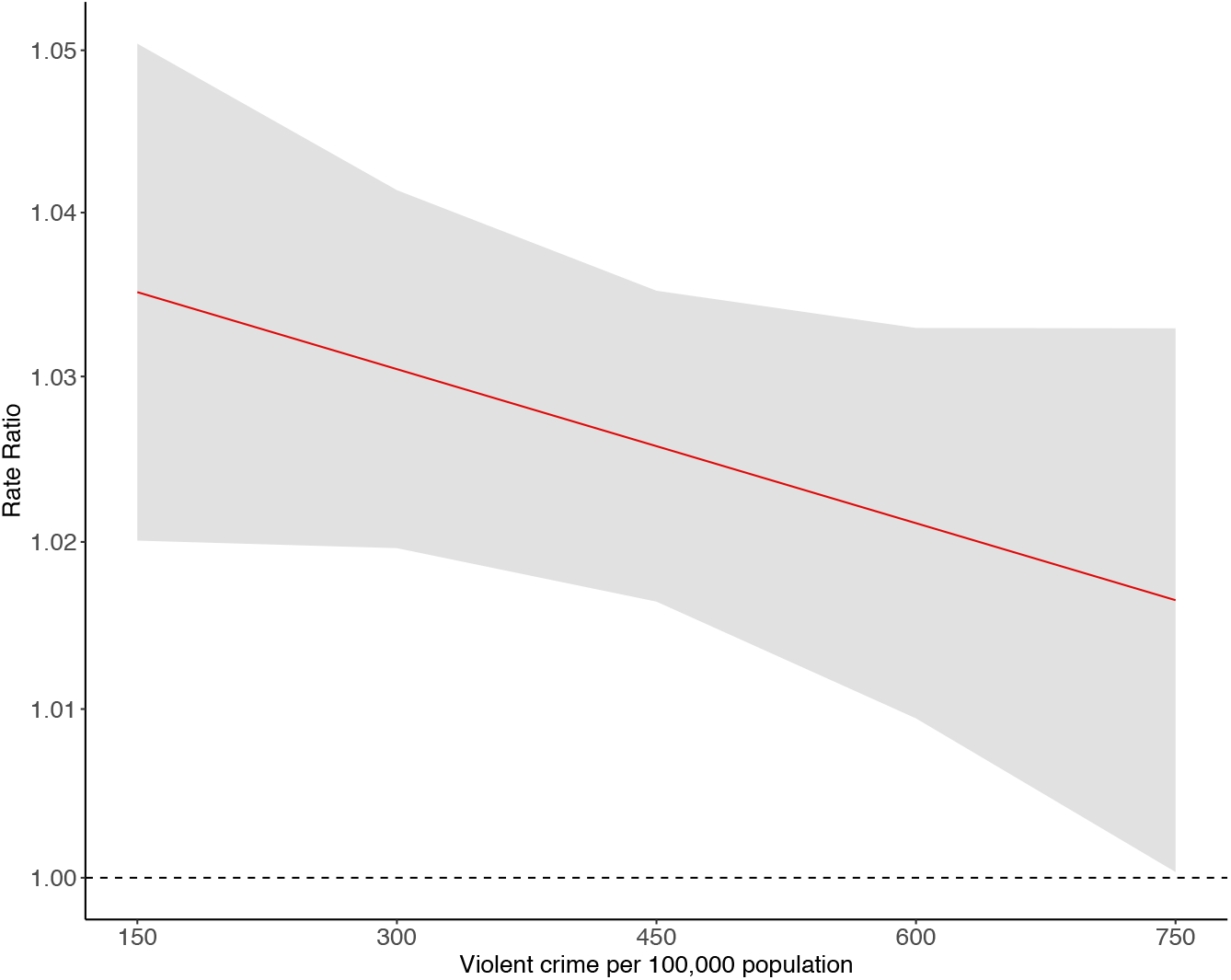
Variation in association between excess firearm purchases and interpersonal firearm violence by states’ baseline violent crime rate. Note. Violent crime rate represents the average state-wide rate from 2014-2018. Rate ratios (not log-transformed) are plotted on the log scale. Wald test for interaction P = 0.15. Rate ratios reflect an increase of 100 purchases per 100,000 population at varying levels of baseline violent crime. We exclude Washington DC, an outlier with a high crime rate; this does not affect results or interpretation.

We do not find significant variation in the association by baseline firearm ownership prevalence, socioeconomic status, racial residential segregation, urbanicity, or social distancing (data not shown). The association did, however, vary by states’ baseline violent crime rate. The relationship between excess purchases and firearm violence was stronger in states with lower pre-COVID-19 rates of violent crime (Figure 2).

Findings were consistent when modeling events (rather than injuries) as the outcome (Supplementary Table 4). We find no evidence of change in the ratio of injuries to events associated with excess purchases (Supplementary Table 5).

Results were robust to defining the exposure as percentage change in purchasing (Supplementary Table 6), excluding DC and events in which children shot another person (Supplementary Table 7), and inclusion of state-specific linear trends (Supplementary Table 8) and all-cause mortality (Supplementary Table 9). Neither 3 nor 6-month leading values of the exposure–defined as the absolute change in purchasing–were associated with firearm violence (Supplementary Table 10). We did, however, find evidence of an association with 6-month, but not 3-month, but not 6-month, leading values of the exposure when defined as the percentage change in purchasing (Supplementary Table 10).

## DISCUSSION

The results of the present study suggest a significant increase in firearm violence in the US associated with the coronavirus pandemic-related surge in firearm purchasing. We estimate a nationwide excess of 2.1 million firearm purchases in March through May 2020. States with greater increases in firearm purchasing were more likely to experience increased rates of firearm violence during this time compared to states with smaller purchasing increases, independent of other effects of the pandemic included in our models. We estimate an almost 8% increase in firearm violence in the US from March through May 2020, or 776 additional injuries, associated with purchasing spikes.

Prior studies have similarly documented an association between firearm violence and spikes in firearm purchasing related to mass shootings and political events. ^11–13^ In a study of handgun purchasing spikes in California following the 2012 presidential election and Sandy Hook shooting, every 100 excess purchases per 100,000 persons was associated with 1.044 times the rate of nonfatal firearm injuries in the year following the spike. ^12^ The magnitude of our estimate is slightly smaller (RR = 1.015)—perhaps indicative of a social distancing-induced decrease in violence in public—but we detect an association immediately following the surge.

The current increase in firearm purchases may be unique, not only in its catalyst but also in its consequences. The risks of increased firearm availability are likely compounded by the myriad effects of the coronavirus pandemic, including widespread increases in anxiety, fear, grief, economic strain, disruptions to daily routines, and racial and economic inequities. ^40,41^ The relationships seen here might not apply to surges arising under other circumstances.

This study lends support to interventions restricting access to firearms. The findings are consistent with individual- and household-level studies that have resulted in recommendations to screen for firearm ownership in healthcare settings, ^42^ support safer firearm use and storage, ^43^ and address contributing risk factors for violence. ^44^ Given the impulsive nature of most firearm violence, and the multiple strains associated with the pandemic, short-term crisis interventions, such as extreme risk protection orders and those involving violence intervention specialists, may be particularly useful during the pandemic.

Further, our findings suggest a need to address perceptions of risk and safety associated with firearm ownership. While we do not have data on people’s motivations for purchasing firearms, anecdotes published in the media suggest that fears for personal safety and possible civil unrest contributed to the current surge in purchasing. ^45^ Prior to the pandemic, firearms, particularly handguns, were more commonly owned for protection against people than for other reasons. ^32^ A large and growing body of literature, including the present study, ties firearms to increased—rather than decreased— risk of firearm injury. ^10,46^ Together, these findings suggest that addressing misperceptions about the health risks and benefits of firearm ownership and improving people’s sense of collective trust and security may reduce the burden of firearm violence.

### Limitations

We cannot infer causality from these observational data. First, though the coronavirus pandemic presents an exogenous shock, our design is subject to confounding insofar as other effects of the pandemic may influence firearm violence through pathways other than changes in purchasing. To mitigate bias, we included all hypothesized and measurable confounders: stay-at-home orders, a measure of compliance with social distancing guidelines, coronavirus cases and deaths, and temperature and precipitation. There may be residual confounding by unmeasured or unmeasurable factors. Second, 6-month leading values of the exposure, when defined as the percentage change in purchasing, were associated with firearm violence rates. This could indicate reverse causation, i.e., that an increase in violence caused an increase in firearm purchasing, that the spike began before March in some states due to earlier effects of the pandemic, or that a third, confounding variable, led states with already high levels of violence to experience greater spikes in purchasing. It is unlikely that our results are explained by these mechanisms, however, because we did not observe an association in the 3 months before March 2020 or when measuring excess purchasing in absolute terms. Despite the limitations of the present study, our estimates are strong and consistent, include evidence of a linear dose-response relationship, and are plausible and consistent with the existing literature. ^47^

There are also data limitations. GVA and NICS data provide imperfect measures of firearm violence and purchasing, respectively. To bias our results, however, there would need to be similarly-timed differential changes across states in GVA or NICS reporting. Disagreement between NICS checks and purchased firearms would most likely result from an increase in multiple-firearm transactions during surges in purchasing, which would introduce a conservative bias in estimates of the number of firearms purchased during surges. Additionally, we have no information on whether the excess firearms acquired were those used in violence. Findings from our study cannot inform relationships at the individual level.

Finally, we measure the short-term impact of changes in purchasing, and we focus narrowly on interpersonal firearm violence; effects may endure over time and extend to other types of firearm violence.

## CONCLUSION

We find that short-term surges in firearm purchasing associated with the coronavirus pandemic are associated with significant increases in interpersonal firearm violence. Our findings are consistent with an extensive literature that documents a link between firearm access and greater risk of firearm violence.

## Data Availability

All data are publicly available. Links and references are provided in the supplement.

## REFERENCES

1. Wintemute GJ. The epidemiology of firearm violence in the twenty-first century United States. Annu Rev Public Health. 2015;36(1):5–19. doi:10.1146/annurev-publhealth-031914-122535

2. Ranney M, Karb R, Ehrlich P, et al. What are the long-term consequences of youth exposure to firearm injury, and how do we prevent them? A scoping review. J Behav Med. 2019;42(4):724–740. doi:10.1007/s10865-019-00035-2

3. Irvin-Erickson Y, Lynch M, Gurvis A, Mohr E, Bai B. A neighborhood-level analysis of the economic impact of gun violence. 2017. Accessed July 1, 2020. https://www.urban.org/research/publication/neighborhood-level-analysis-economic-impact-gun-violence/view/full_report

4. Miller M. Firearm availability and suicide, homicide, and unintentional firearm deaths among women. J Urban Health Bull N Y Acad Med. 2002;79(1):26–38. doi:10.1093/jurban/79.1.26

5. Miller M, Azrael D, Hemenway D. Firearm availability and unintentional firearm deaths, suicide, and homicide among 5–14 year olds: J Trauma Acute Care Surg. 2002;52(2):267–275. doi:10.1097/00005373-200202000-00011

6. Miller M, Azrael D, Hemenway D. Rates of household firearm ownership and homicide across us regions and states, 1988–1997. Am J Public Health. 2002;92(12):1988–1993. doi:10.2105/AJPH.92.12.1988

7. Kellermann AL, Rivara FP, Somes G, et al. Suicide in the home in relation to gun ownership. N Engl J Med. 1992;327(7):467–472. doi:10.1056/nejm199208133270705

8. Kellermann AL, Rivara FP, Rushforth NB, et al. Gun ownership as a risk factor for homicide in the home. N Engl J Med. 1993;329(15):1084–1091. doi:10.1056/nejm199310073291506

9. Wintemute GJ, Parham CA, Beaumont JJ, Wright M, Drake C. Mortality among recent purchasers of handguns. N Engl J Med. 1999;341(21):1583–1589. doi:10.1056/NEJM199911183412106

10. Studdert DM, Zhang Y, Swanson SA, et al. Handgun ownership and suicide in California. N Engl J Med. 2020;382(23):2220–2229. doi:10.1056/NEJMsa1916744

11. Studdert DM, Zhang Y, Rodden JA, Hyndman RJ, Wintemute GJ. Handgun acquisitions in California after two mass shootings. Ann Intern Med. 2017;166(10):698. doi:10.7326/M16-1574

12. Laqueur HS, Kagawa RMC, McCort CD, Pallin R, Wintemute G. The impact of spikes in handgun acquisitions on firearm-related harms. Inj Epidemiol. 2019;6(1):35. doi:10.1186/s40621-019-0212-0

13. Levine PB, McKnight R. Firearms and accidental deaths: Evidence from the aftermath of the Sandy Hook school shooting. Science. 2017;358(6368):1324–1328. doi:10.1126/science.aan8179

14. COVID-19 United States Cases by County. Johns Hopkins Coronavirus Resource Center. Accessed June 9, 2020. https://coronavirus.jhu.edu/us-map

15. NICS Firearm Checks: Month/Year by State/Type. Federal Bureau of Investigation. Accessed June 2, 2020. https://www.fbi.gov/file-repository/nics_firearm_checks_-_month_year_by_state_type.pdf/view

16. Gun Violence Archive incident search. https://www.gunviolencearchive.org/query

17. Conner A, Azrael D, Lyons VH, Barber C, Miller M. Validating the National Violent Death Reporting System as a source of data on fatal shootings of civilians by law enforcement officers. Am J Public Health. 2019;109(4):578–584. doi:10.2105/AJPH.2018.304904

18. Kim D. Social determinants of health in relation to firearm-related homicides in the United States: A nationwide multilevel cross-sectional study. Brohi K, ed. PLOS Med. 2019;16(12):e1002978. doi:10.1371/journal.pmed.1002978

19. Booty M, O’Dwyer J, Webster D, McCourt A, Crifasi C. Describing a “mass shooting”: the role of databases in understanding burden. Inj Epidemiol. 2019;6(1):47. doi:10.1186/s40621-019-0226-7

20. Klassen AB, Marshall M, Dai M, Mann NC, Sztajnkrycer MD. Emergency medical services response to mass shooting and active shooter incidents, United States, 2014–2015. Prehosp Emerg Care. 2019;23(2):159–166. doi:10.1080/10903127.2018.1484970

21. Donnelly L, McLanahan S, Brooks-Gunn J, James S, Rouhani S. Disparities in adolescents’ recent exposure to local gun violence: linking incident-level crime data to a population-based panel study. Presented at the: 2018 annual meeting of the Population Association of America; April 26, 2018; Denver, CO.

22. Mervosh S, Lee JC, Gamio L, Popovich N, Matthews AL. See which states are reopening and which are still shut down. The New York Times. https://www.nytimes.com/interactive/2020/us/states-reopen-map-coronavirus.html. Published April 20, 2020. Accessed June 4, 2020.

23. Mervosh S, Lee JC, Gamio L, Popovich N. See how all 50 states are reopening (and closing again). The New York Times. https://www.nytimes.com/interactive/2020/us/states-reopen-map-coronavirus.html. Published June 4, 2020. Accessed June 4, 2020.

24. COVID-19 - Mobility Trends Reports. Apple. Accessed June 14, 2020. https://www.apple.com/covid19/mobility

25. PRISM Climate Group, Oregon State University, Time series values for multiple locations. Accessed June 4, 2020. https://prism.oregonstate.edu/explorer/bulk.php

26. Hyndman RJ, Khandakar Y. Automatic time series forecasting: the forecast package for R. J Stat Softw. 2008;27(3). doi:10.18637/jss.v027.i03

27. Ljung GM, Box GEP. On a measure of lack of fit in time series models. Biometrika. 1978;65(2):297–303. doi:10.1093/biomet/65.2.297

28. Benjamini Y, Hochberg Y. Controlling the false discovery rate: a practical and powerful approach to multiple testing. J R Stat Soc Ser B Methodol. 1995;57(1):289–300. doi:10.1111/j.2517-6161.1995.tb02031.x

29. Allison PD, Waterman RP. 7. Fixed-effects negative binomial regression models. Sociol Methodol. 2002;32(1):247–265. doi:10.1111/1467-9531.00117

30. Fixed Effects Models for Count Data. In: Fixed Effects Regression Models. SAGE Publications, Inc.; 2009:49–69. doi:10.4135/9781412993869.d20

31. Jewell NP. Statistics for Epidemiology. Chapman & Hall/CRC; 2003. Accessed July 1, 2020. https://public.ebookcentral.proquest.com/choice/publicfullrecord.aspx?p=199157

32. Azrael D, Hepburn L, Hemenway D, Miller M. The stock and flow of U.S. firearms: results from the 2015 National Firearms Survey. RSF. 2017;3(5):38–57. doi:10.7758/rsf.2017.3.5.02

33. Fatal Injury Data. Web-based Injury Statistics Query and Reporting System (WISQARS). Centers for Disease Control and Prevention. Published July 1, 2020. Accessed July 1, 2020. https://www.cdc.gov/injury/wisqars/fatal.html

34. Rankings Data & Documentation. County health rankings & roadmaps. Accessed June 14, 2020. https://www.countyhealthrankings.org/explore-health-rankings/rankings-data-documentation

35. Massey DS, Denton NA. The dimensions of residential segregation. Soc Forces. 1988;67(2):281. doi:10.2307/2579183

36. Rankings Data & Documentation with data from the Census. County Health Rankings & Roadmaps. Accessed June 14, 2020. https://www.countyhealthrankings.org/explore-health-rankings/rankings-data-documentation

37. Federal Bureau of Investigation. Uniform Crime Reporting. Accessed July 1, 2020. https://crime-data-explorer.fr.cloud.gov/downloads-and-docs

38. Keasey CB, Sales BD. Children’s conception of intentionality and the criminal law. Psychol Leg Process. Published online 1977. https://www.ncjrs.gov/App/Publications/abstract.aspx?ID=51499

39. Wing C, Simon K, Bello-Gomez RA. Designing difference in difference studies: best practices for public health policy research. Annu Rev Public Health. 2018;39(1):453–469. doi:10.1146/annurev-publhealth-040617-013507

40. Torales J, O’Higgins M, Castaldelli-Maia JM, Ventriglio A. The outbreak of COVID-19 coronavirus and its impact on global mental health. Int J Soc Psychiatry. 2020;66(4):317–320. doi:10.1177/0020764020915212

41. Okonkwo NE, Aguwa UT, Jang M, et al. COVID-19 and the US response: accelerating health inequities. BMJ Evid-Based Med. Published online June 3, 2020:bmjebm-2020-111426. doi:10.1136/bmjebm-2020-111426

42. Pallin R, Spitzer SA, Ranney ML, Betz ME, Wintemute GJ. Preventing firearm-related death and injury. Ann Intern Med. 2019;170(11):ITC81. doi:10.7326/AITC201906040

43. Shenassa ED, Rogers ML, Spalding KL, Roberts MB. Safer storage of firearms at home and risk of suicide: a study of protective factors in a nationally representative sample. J Epidemiol Community Health. 2004;58(10):841–848. doi:10.1136/jech.2003.017343

44. Kagawa RMC, Stewart S, Wright MA, et al. Association of prior convictions for driving under the influence with risk of subsequent arrest for violent crimes among handgun purchasers. JAMA Intern Med. 2020;180(1):35. doi:10.1001/jamainternmed.2019.4491

45. Collins K, Yaffe-Bellany D. About 2 million guns were sold in the U.S. as virus fears spread. The New York Times. https://www.nytimes.com/interactive/2020/04/01/business/coronavirus-gun-sales.html. Published April 2, 2020.

46. Anglemyer A, Horvath T, Rutherford G. The accessibility of firearms and risk for suicide and homicide victimization among household members: a systematic review and meta-analysis. Ann Intern Med. 2014;160(2):101–110. doi:10.7326/m13-1301

47. Hill AB. The environment and disease: association or causation? Proc R Soc Med. 1965;58(5):295–300.

